# The cascading impacts of attacks on health in Syria: A qualitative study of health system and community impacts

**DOI:** 10.1101/2023.12.18.23300154

**Authors:** Rohini J. Haar, Diana Rayes, Hannah Tappis, Leonard Rubenstein, Anas Rihawi, Mohamed Hamze, Naser Almhawish, Reham Wais, Hesham Alahmad, Ryan Burbach, Aula Abbara

## Abstract

**Introduction:** Syria has experienced over a decade of armed conflict, characterized by targeted violence against healthcare. The impacts of these attacks have resulted in both direct and indirect attacks on the health system and reverberating effects on local communities. This study aims to explore the perspectives of health workers based in northern Syria who have experienced such attacks on health to understand the impacts on the health system as well as communities served.

**Methods:** In-depth interviews were conducted with health workers in the northern regions of Syria where attacks on health have been frequent. Participants were identified using purposive and snowball sampling. Interviews were coded and analyzed using the Framework Method. Our inductive and deductive codes aligned closely with the WHO Health System Building Blocks framework, and we therefore integrated this framing into the presentation of findings. We actively sought to include female and non-physician health workers as both groups have been under-represented in previous research in northern Syria.

**Results:** A total of 40 health workers (32.5% female, 77.5% non-physicians) who experienced attacks in northern Syria between 2013 and 2020 participated in interviews in 2020-2021. Participants characterized attacks on healthcare as frequent, persistent over years and strategically targeted. The attacks had both direct and indirect impacts on the health system and consequently the wider health of the community. For the health system, participants noted compounded impacts on the delivery of care, health system governance, and challenges to financing, workforce, and infrastructure. Reconstructing health facilities or planning services in the aftermath of attacks on health was challenging due to poor health system governance and resource challenges. These impacts had ripple effects on the health of the community, particularly the most vulnerable.

**Discussion:** The impacts of attacks on health in Syria are multiple, with both short- and long-term consequences for the health system(s) across Syria as well as the health of communities in these respective areas. Though such attacks against healthcare are illegal under international humanitarian law, this and other legal frameworks have led to little accountability in the face of such attacks both in Syria and elsewhere. Characterizing their impacts is essential to improving our understanding of the consequences of attacks as a public health issue and supporting protection and advocacy efforts.

## I. Introduction

Violence against healthcare in conflict settings is a tragic yet common occurrence in recent years, with devastating consequences on healthcare providers, patients and the entire health system.[1-3] These attacks can be characterized by direct violence, such as airstrikes of hospitals and assaults of health workers or patients. They also include arrests, threats, intimidation, and interference with health services, all of which violate International Humanitarian Law in armed conflict.[4] Indirect impacts include the effects on healthcare access due to damaged healthcare facilities and infrastructure, the forced exodus of healthcare workers (who are often the targets of direct attacks), underfunding and a reluctance for patients to seek healthcare as they perceive health facilities to be unsafe.[6] Numerous and diverse attacks on healthcare have been recorded over the past two decades, with the most frequent violations reported in Afghanistan, Central African Republic, Colombia, Ethiopia, Myanmar, Palestine, South Sudan, Syria, Yemen, and, most recently, Ukraine and Sudan, perpetrated both by internal and external armed actors and militaries.[1,5] Documentation of these attacks is critical, and mandated by the UN Security Council (Resolution 2286).[6] In 2022 alone, the Safeguarding Health in Conflict Coalition and Insecurity Insight reported nearly 2,000 incidents of attacks on healthcare facilities and personnel in 32 countries, with 232 health workers killed and hundreds more kidnapped and arrested.[3]

Syria’s brutal conflict began after peaceful uprisings in March 2011 were violently and disproportionately suppressed by the Syrian government with significant disruption to its health system.[7,8] The criminalization of the provision of medical care to those considered opposed to the government and the direct attacks and disruption to healthcare began soon after.[9] As the conflict progressed, hundreds of aerial bombardments of healthcare facilities were recorded over the years, mostly in parts of the countries opposed to the Syrian government, along with other forms of violence against healthcare including the interruption of healthcare access and supplies.[10]

Many involved parties, including the governments of Syria, Russia, the United States and multiple non-state armed groups including the Islamic State, al-Nusra Front and others, are implicated in such violence, however the vast majority of strikes on health were conducted by Syrian government and aligned Russian forces in opposition-controlled areas, primarily the cities of Idlib and northern Aleppo.[9-11]. According to Physicians for Human Rights, between 2011 and March 2022, there have been more than 601 airstrikes on at least 222 health care facilities, with evidence that these strikes were intentional and targeted.[2,7,11-13] Over 900 health workers have been killed directly in the conflict, many in the course of their work and untold more have suffered from physical and psychological injuries.[13,14] The number of patients affected is unknown but includes those with immediate injuries as well as many who avoided seeking healthcare, waited too long, were afraid to report stigmatized violence, or were deprived of regular treatment or preventative services; the impacts on this latter group are much harder to quantify.[13,15] Attacks on healthcare in Syria has also be an important driver of forced displacement within and outside of Syria’s borders as well as a key driver of the forced exodus of health workers.[7]

The evidence available suggests that attacks on healthcare have wide-ranging impacts on the delivery of healthcare, including long-term health consequences, economic losses, and an erosion of trust in healthcare systems.[7-10]. However, a detailed exploration of these impacts and their nature on the health system in Syria and elsewhere are limited, as are studies of the experiences of health workers subject to attacks.[16-20] The perspectives of health workers who have survived attacks is critical for gaining a deeper understanding of the impacts of violence on the many components of the health system in Syria.

This study aims to explore the complex and compounding impacts of violence on the health system, including its effects on the availability and accessibility of care, selected health outcomes and the long-term stability of health resources. Employing a health systems approach facilitated organizing impacts within a structured framework that would facilitate a deeper comprehension of the impacts, both directly and in aggregate.[21,22]

## II. Methods

### Research design

The study was conducted by researchers at the University of California, Berkeley and Johns Hopkins University in partnership with two humanitarian organizations working directly on the humanitarian health response inside Syria: the Syrian American Medical Society (SAMS) and Assistance Coordination Unit (ACU). We used a qualitative research design with in-depth, semi-structured interviews with Syrian healthcare workers who had experienced attacks on healthcare during the conflict. We developed the interview guide based on previous research on attacks on health carried out by members of the study team in Syria, Myanmar and Colombia and in coordination with our local partners with a content analysis orientation.[16,17,25] The overall research question guiding this study was: What are health worker perceptions of the impact of violence on healthcare based on their personal experiences and knowledge of attacks?

### Partner organizations

SAMS was established in 1998 and their humanitarian arm has provided health and humanitarian care in Syria and in Syrian refugee hosting countries since the onset of the uprisings in 2011.[23] ACU, established in 2013, manages the Early Warning Alert and Response Network (EWARN) surveillance program as well as several other key health and Water Sanitation and Hygiene (WASH) monitoring and information systems across areas in Syria which are outside of government control. Both organizations maintain ongoing programs in Syria and have established networks of health workers in Syria.[24] Partnering at the conceptualization, execution and analysis phases of the study was intentional and aimed to support co-design of the study from researchers who themselves had experience of attacks on health in Syria.

### Setting and participant selection

Study participants were recruited through purposive and snowball sampling though SAMS’ and ACU’s professional networks. This approach enabled us to promote interviews with female and non-physician health workers as both groups are under-represented in conflict research. We defined the term health worker broadly to include physicians as well as midwives, nurses, pharmacists, dentists as well as technicians (e.g. anesthetic, dialysis) and those who held administrative or management roles. Other inclusion criteria included personal experiences of attacks on healthcare in northern Syria between 2011 and 2020, ability and willingness to consent and an internet connection.

Participants were included from governorates within northwest Syria including parts of Aleppo, Idlib, Hama, and Homs governorates and to a lesser extent, northeast Syria including parts of Al-Hasakah, Aleppo (Menbij and Ain Al Arab), Deir Ez Zor, and Raqqa. These governorates were selected based on the high frequency of attacks in these areas as well as the accessibility, connectivity and security of health workers available for interviews (Figure 1).[8,13,14]

**Figure 1.**
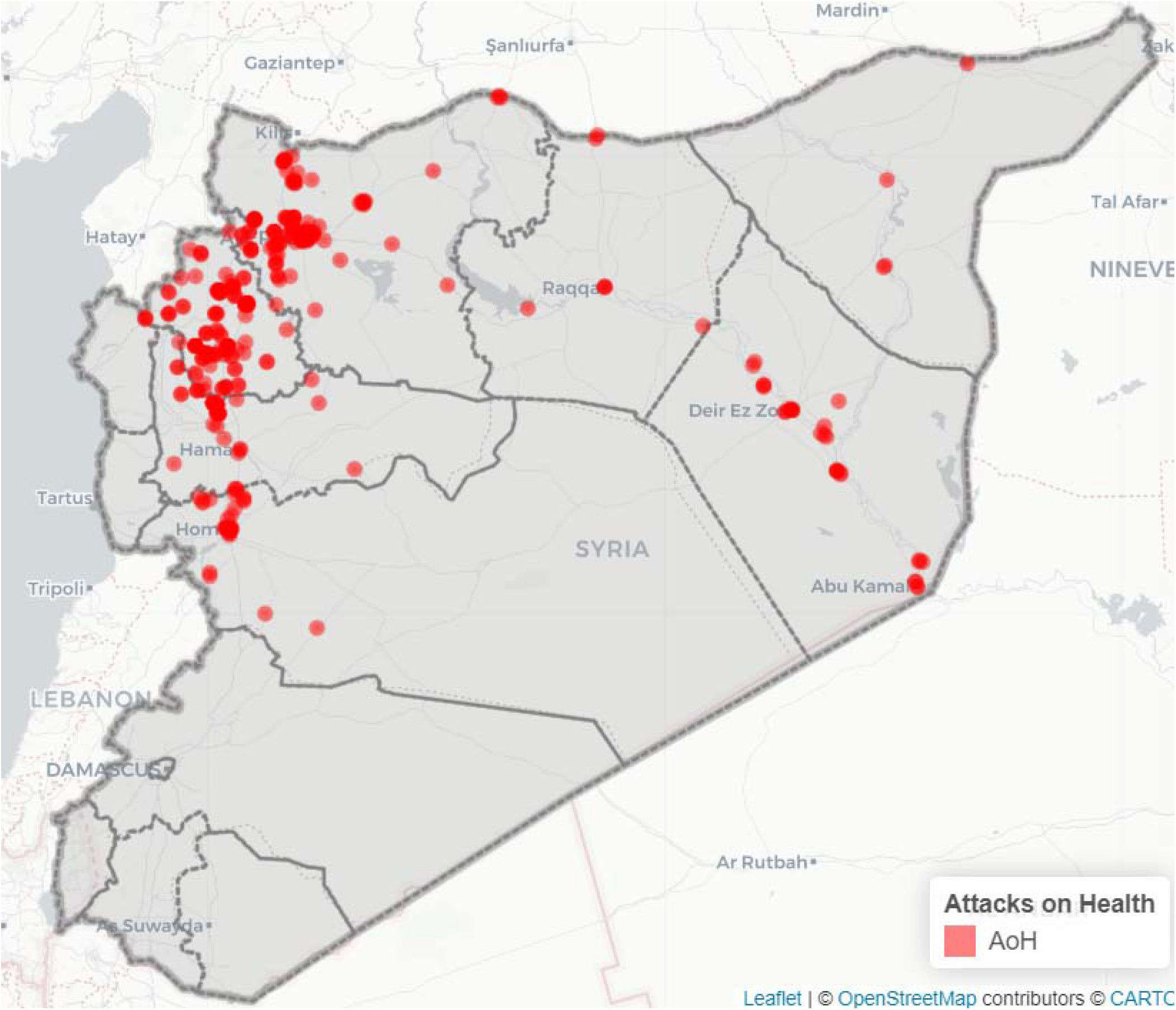
Map of attacks reported in northern Syria: source PHR and SAMS]

### Data collection

All interviews were conducted in Arabic by Syrian-American team members (DR and AR) using Zoom (version 5.0) for accessibility and safety reasons. Both interviewers were fluent in Syrian Arabic, experienced in conflict research and practiced reflexivity throughout the process. Most interviews were conducted by a female researcher (DR) to encourage female participation. Following oral consent, participants were asked to describe their roles and responsibilities as well as their experiences of the Syrian conflict, particularly violence against healthcare. After the interview, participants were asked to refer additional potential participants to the research team. Data collection was terminated once participants were no longer presenting major additional concepts and thematic saturation was achieved.

Most interviews were audio-recorded for the transcription of interviews and in order to maintain the accuracy of information shared. In a few cases, handwritten notes were taken instead of recording based on participant comfort and preference. Interviews were transcribed in Arabic and translated to English by native Arabic speakers for coding and analysis. We extracted participants’ demographic data including profession, location and gender and specific incidents of violence they described. Data were analyzed using Dedoose (version 9.0) qualitative software.

### Data analysis

We analyzed the translated English transcripts using the Framework Method, which is well-suited to analyze health research using a multi-disciplinary approach.[26] One key distinction of this approach is that it permits both inductive and deductive approaches to thematic analysis. We followed the steps proposed by the Framework Method including familiarization with transcripts, initial coding, developing an analytical framework (set of codes that are mutually agreed upon), and charting data into a framework matrix. A codebook was developed based on the broader study objective of exploring the short- and long-term impacts of attacks on a) healthcare facilities, b) health services, c) community health, d) health workers, and e) the health system.

We found that our deductive and inductive codes aligned closely with the WHO Health System Building Blocks framework and therefore integrated this framing into our analysis.[27] We focused on five out of the six existing WHO Health System Building Blocks: 1) service delivery 2) health workforce, (3) essential medicines and infrastructure, (4) governance, (5) financing. Consistent with recent guidance on adapting health system components for context-specific research, we replaced the sixth building block (Health Information Systems) with the domain of (6) security and protection, as these concepts are particularly relevant to the humanitarian context.[28] We approach governance as it relates to health in a similar vein to WHO in terms of stewardship,’ oversight, regulation, incentives, and accountability.[29] The connection between the building blocks and the clinical experience varies which could influence the analysis given impacts are examined via the perspective of health workers in this study.

Interviews were both deductively and inductively coded by at least two individual coders (DR, AR, RH, HT) and compared and discussed for discrepancies. Coding and subsequent interview results were discussed iteratively among the qualitative research team on a regular basis, identifying potential themes of interest and areas for further exploration. Themes were developed based on the perceptions of health workers regarding the various impacts of violence against healthcare as well as in alignment with the WHO Health System Building Blocks framework. We identified key patterns, and correlations among the responses and highlighted quotations germane to the analysis. Emerging findings from the qualitative data were also discussed with the larger study group in order to probe deeper, ensure consistency, and explore any incongruous data.

### Ethical Considerations

The University of California, Berkeley’s Human Subjects Protection Program reviewed and approved this study (Protocol # 2020-03-13069). The researchers also consulted with and received approval from the Idlib Health Directorate and other local leadership in lieu of a formal ethical review board in Syria.

Comprehensive informed consent procedures took place at the start of each interview. Participants were able to terminate the interview at any time and to withdraw their participation until the time at which analysis was started. Given that some topics covered may be sensitive or cause distress, a reference to a mental health professional was available should this be needed by the participants.

## III. Results

Between June 2020 and November 2021, a total of 40 health workers participated in remote in-depth interviews. Participants included 27 males (67.5%) and 13 females (32.5%) and represented a variety of health professions, including nurses (25%), physicians (including dentists, surgeons, and medical doctors) (22.5%), technicians (laboratory, surgical assistants, and anesthetics) (15%), health administrators (10%), first responders (10%), pharmacists (5%), medical students (7.5%), a midwife (2.5%), and a psychologist (2.5%). All participants experienced attacks on healthcare facilities between 2013 and 2020, most of them experienced more than one.

**Table 1.** Participant characteristics.

| Total Participants | 40 | N (%) | Total Participants | 40 | N (%) |
| --- | --- | --- | --- | --- | --- |
| **Location** | Aleppo | 12 (30) | **Profession** | Administrator | 4 (10) |
|  | Ghouta | 3 (7.5) |  | First Responder | 4 (10) |
|  | Hama | 2 (5) |  | Midwife | 1 (2.5) |
|  | Idlib | 18 (45) |  | Nurse | 10 (25) |
|  | Kafranbel | 1 (2.5) |  | Pharmacist | 2 (5) |
|  | Raqqa | 4 (10) |  | Physician | 9 (22.5) |
| **Age Range** | 20-29 | 14 (35) |  | Psychologist | 1 (2.5) |
|  | 30-39 | 12 (30) |  | Student | 3 (7.5) |
|  | 40-49 | 2 (5) |  | Technician | 6 (15) |
|  | 50-59 | 5 (12.5) | **Gender** | Female | 13 (32.5) |
|  | ≥60 | 2 (5) |  | Male | 27 (67.5) |
|  | Unknown/not shared | 5 (12.5) |  |  |  |

Participants were mainly based in northwest Syria, in Idlib and Aleppo governorates, followed by a smaller number from northeastern Syria, including Raqqa and central Syria, including Eastern Ghouta, and Hama. They worked in facilities experiencing attacks including general hospitals (46%), surgical centers (33%), primary care centers (13%), and specialty hospitals (8%). Numerous medical facilities, clinics, and ambulances were named as sites where health workers were working or experienced attacks. Almost all health workers had experienced more than one attack and had been displaced several times over the course of the conflict. All the participants were still working in the health sector in some capacity; however, at least 5 of the participants were not working in the profession that they had initially received training in.

Health workers described that violence against healthcare in Syria resulted in intersectional and compounded impacts which affect all parts of the health system. Adapted from the WHO Health System Building Blocks framework, we outline both direct and indirect impacts under the following themes: 1) severe constraints on *health service delivery*, 2) challenges in health *workforce* retention and morale, 3) compounding loss in *essential medicines and infrastructure*, 4) weakened *health governance*, 5) uncertainties in health *financing*, and 6) compromised *security and protection* [27,28]. Collectively, these health system impacts had cascading impacts on community health.

### Health System Impacts

#### 1) Severe constraints on health service delivery

All participants emphasized that the quantity and quality of health service delivery was affected by attacks on healthcare in both the short- and the long-term. This was due to a combination of direct damage to healthcare facilities, as well as increased health needs and demand on facilities due to the direct and indirect impacts of the conflict. At times of an acute increase in violence leading to forced displacement, participants reported that there could be a sudden and significant population growth in some areas, leading to a spike in demand.

##### Irreparable damage to health facilities

One participant observed that, “if this destroyed hospital was providing healthcare services to a particular area of half a million people, then that half a million people would be without healthcare for a month. They may also have to go to other areas and their diagnosis may be delayed, as well as having complications and many other problems.” (Administrator) Participants noted disparities between urban and rural facilities; in rural areas, when attacks on health facilities occurred resulting in closures, those communities had more limited options for alternative healthcare.

Even when health facilities were re-opened, all “services were not always fully reinstated” stated a participant who highlighted this reality with an example of a pediatric unit that remained closed in order to prioritize the reopening of the trauma unit (Physician). Chronic and preventative care, as well as care requiring specialized or high-resource equipment, took longer to reinstate. Another participant noted that early discharges of admitted patients were frequent when the facility was felt to be at higher risk of being attacked or had been attacked recently. In many cases, attacks also impacted quality of care. “If we want to give each patient five minutes, the patient does not [get what they need] but these are the available capabilities. (Lab Technician).

##### Ripple effects on nearby facilities

The reduction in services had ripple effects on other health facilities in the patient catchment area and their ability to meet demand, often overburdening still-functioning or partially functioning facilities. Several participants described that patients were transferred to or advised to go to alternative facilities, especially those outside city centers, when attacks occurred. One participant described the scale of redistribution of patient loads: “The hospital has recently become the one to serve the entire south of Idlib. If you know southern Idlib, all hospitals were bombed before us, then all hospitals were stopped, so [our] hospital received patients from Ariha, rural Ariha, and southern rural Idlib; even northern rural Hama and southern rural Aleppo…. The workload - it was intense.” (Nurse)

##### Irretrievable loss of specialized services

Specialized services were particularly affected if provider facilities were damaged; this included services for chronic conditions, including cancer, kidney failure (dialysis) and mental health conditions. Mental health needs among the community increased significantly due to the conflict and were noted by participants to be unmet due to the lack of mental health specialists (which also pre-dated the conflict), but was exacerbated by attacks, and the fear among patients of visiting hospitals. Cancer services were also noted to be a particularly scarce resource in these areas.

A participant described the difficult decisions that patients need to make, “For example, cancer patients now have two options: either to head towards Damascus or towards Turkey” (Lab Technician) which is not always an option for all patients. This is particularly due to social factors which, as one participant noted, particularly affected widows or people living with disability because they could not travel as easily. (Physician)

#### 2) Challenges in health workforce retention and morale

Attacks on healthcare were described to result in a precipitous drop in the availability of workers through forced displacement, a lack of training opportunities, among other reasons.

The lack of trained health workers had catastrophic effects on some services. One participant illustrated this experience:

> “We still did not have skilled general practitioners who knew how to use the triage system and how to treat 90% of the common diseases in the community… The number of gynecologists was also very, very few compared to the number of women in the community… That is why a lot of complications and mortality were occurring. This is what we have… there are not enough physicians… What can we do?!” (Physician)

##### Displacement and unemployment

Many health workers experienced forced displacement, often more than once. The wide-ranging impacts of this were noted to affect the quality of care for patients as well as the work experiences of health workers. Several participants described disruption to their ability to develop long-term rapports with their patients and strong relationships with their colleagues. A participant also noted the critical importance of establishing a long-term relationship with patients, particularly in settings where clinical records are absent, poorly kept or destroyed. Other participants described that being unable to continue working with well-established and familiar colleagues was challenging. Some also described a different culture in different cities in Syria or that they felt like outsiders. “In Idlib, the culture is different. They call us *nazeheen* (displaced) – but we did not leave by our own choice” said one participant who had been displaced more than 15 times. (Nurse)

An important but paradoxical workforce challenge was that in the wake of an attack, despite increased needs for services, health workers could become unemployed if funding to a facility was interrupted or redirected, which often occurred following attacks. One participant said that, “Physician retention is difficult as the hospital is rebuilding and [health workers] need to find work in the meantime and then come back.” (Medical Technician). This resulted in unstable and inconsistent work as a direct result of attacks which severely damaged health facilities. Another participant explained that “health workers live in an ambiguous future—[the] near future and far future are both not clear.” (Physician)

##### Skills and training

Staff shortages resulting from the targeting and the exodus of health workers has led to an increase in task-shifting, a phenomenon which was uncommon before the conflict. Health workers were forced to take on roles that they were not prepared or trained for, causing undue stress among health workers and their colleagues, and potentially leading to suboptimal quality of care for patients. One participant highlighted this gap, particularly in relation to maternal health as there were too few obstetricians, stating, “..and midwives have only basic training so [we] have far more home births and maternal mortality than before the war.” (Physician)

##### Resistance and solidarity gave strength

Despite the challenges noted by participants, there were also many examples of coping, survival, resistance and solidarity among health workers. Participants stayed in Syria despite the risk it posed to their lives and their well-being and continued to provide services, motivated by a duty towards their respective communities and wanting to meet ongoing needs. However, many participants rejected narratives of “hero” or “victim,” rather just feeling an obligation to help. One participant described this sentiment as, “The feeling that dominates me most is that there is a wounded person in need of rescue. He’ll die if I don’t do my job, its possibility is 100%, whereas the probability of my death in targeting is 5%, the feeling of rescue always prevails over me.” (Physician).

Solidarity both with the Syrian cause as well as with colleagues and staff also played a large role in motivations to continue work. Many health workers described that they communicate via groups on phone messaging applications to stay connected with colleagues, support patients, share stories and find mutual strength. Some participants worked in Syria and visited their families weekly or monthly in Turkey, who lived there for safety reasons, finding balance between work and family. Participants also described feelings of resistance against the Syrian regime that drove them to continue working. One participant described that “For me, I absolutely refuse to leave Syria. In my opinion we must do anything that can support the Syrian revolution, God willing.” (Administrator)

#### 3) Compounding loss of essential medicines and infrastructure

##### Damaged infrastructure

Participants discussed the extensive destruction of infrastructure in Syria due to attacks, including hospitals, mobile clinics, ambulances and supply trucks. Damaged facilities were assessed in the aftermath of attacks and health workers supported the repair efforts and continued to see patients. However, compounded damage from multiple attacks made reconstruction efforts more costly. One participant noted that, “If it was minor damage, [we] can build back in 2-3 weeks. But with severe damage, especially cumulative— it would be hard to ever build back.” (Physician) The perceived high risk of future attacks often made funders and organizations reluctant to rebuild health infrastructure.

##### Scarcity in medical resources

Participants reported that interrupted medical supplies and medications presented challenges for the health system. This was not only due to the direct effects of attacks on health on pharmacy stocks but also damage to transport infrastructure and relevant hospital infrastructure such as electricity, particularly affecting medications that required refrigeration. Even if classes of medications were available, specialist medications, or specific dosages and formulations were scarce. As a result of severe shortages, the cost of medications rose sharply, leaving many medications out of reach for many people who may have needed them. While health workers described a range of strategies to fill the gaps in medication coverage, including free pharmacies within facilities and negotiated discounts on medications coordinated among pharmacists, many patients were still unable to access their medications regularly, also out of fear of visiting health facilities in order to do so. One participant explained the downstream impacts of attacks on healthcare on medication shortages: “Many people cannot buy medicine because it is so expensive…Many people are displaced and in need, have left their jobs, lands, and livelihoods, and yet they are compelled to buy this drug.’ (Physician)

#### 4) Weakened health governance

##### Hindered effectiveness

Participants shared some glimpses of their experience with health governance in northern Syria. Several of them described that the Syrian government’s violence against healthcare includes deliberately undermining the establishment and effectiveness of local health governance and leadership in northern Syria. Attacks on healthcare have thus left both local and international humanitarian organizations to fill this gap. However, given the enormous challenges of building a new health system in the setting of war as well as targeted violence and the lack of executive authority, health governance actors in northern Syria faced numerous challenges, some of which participants touched on in the interviews. Challenges included managing security, ensuring that health facilities stayed open, staffed, and equipped, equitable and efficient policy making and procedures for rebuilding, and prioritizing limited resources. One participant noted that “the main infection in society [is that] there is no direct health authority, there is no security authority … Unfortunately, things have become random, …, there is no regulation, and there is no one to impose curfews, stabilize markets, and regulate the work.” (Administrator)

##### Limited planning and coordination

Participants noted that one of the key challenges for local governance actors (which include health directorates and humanitarian organizations) in northern Syria is the rehabilitation of health facilities damaged by attacks on health. Participants noted that they, as health workers, were also required to make challenging decisions, for example, around fortifications, locations of health facilities to minimize the risk of attacks, and where to prioritize resources. Several noted that prioritizing protection from attacks had trade-offs: more secure locations could restrict access. Participants described that health facilities were increasingly fragmented or siloed in some parts of opposition-controlled Syria. While peacetime structures are often built in central and accessible areas of communities, new health facilities were built away from the centers of town to protect civilians in case they were targeted. One participant described why: “Hospitals cost a lot to build. And one bomb can destroy the whole thing, so it is better to build more smaller ones. We deliberately increased the medical points in order to reduce the damage.” (Physician) Fragmentation of the health system(s) often resulted in inequities in healthcare services for different communities. For instance, one participant stated that, “physicians and health care workers were not distributed evenly—[there were] more in safer areas, and fewer in dangerous areas.” (Physician).

When rebuilt, services were also divided into separate facilities to ensure that a strike on one facility would not affect too many people or services. While this was effective for keeping some services accessible, it made holistic and coordinated medical care for patients challenging.

#### 5) Uncertainties in health financing

Both the direct airstrikes on health facilities and the indirect impact on services were associated with a high cost of healthcare system(s) in northern Syria, as referenced above regarding the increasing cause of medications. According to participants, increasingly limited financial resources over the protracted conflict resulted in tensions around broader medical resource allocation among the different actors and communities, as well as significant dependency on unstable donor funds and international aid (often restricted because of anti-terrorism regulations), and frequent interruptions of funding streams to health service organizations, facilities and staff. In addition, participants perceived that funders were reportedly reluctant to fund health facilities or services at high risk of repeat attacks.

Several participants described challenges with resource allocation both at administrative and clinical levels. Administrators noted that with restricted funds and ongoing losses of infrastructure, rebuilding facilities was not a priority, especially when the risk of re-attack was high. One participant noted that, “The cost of establishing … a hospital was more than two or three million dollars… It is too costly… And in the end, after you pay all these costs, the regime with just one missile can destroy the entire hospital.” (Physician). One participant noted that the persistence of attacks over years also limited healthcare actors and donors from investing in establishing more sophisticated specialized services for tertiary care, concerned that health facilities continued to be targeted.

##### Interrupted funding streams

Unstable donor funds, top-down decisions on funding priorities (without considerations of local needs), and political or technical restrictions on funding resulted in unreliable and frequently interrupted funding streams for the health system. Several participants highlighted that salary delays caused by shifts in donor priorities, coupled with the absence of health and life insurance for workers, demotivated healthcare professionals from staying in perilous conditions and prompted them to contemplate migration. When funding was reduced or interrupted, and health workers were unemployed, participants described how this negatively impacted their livelihoods: “costs actually go up—distance to work from a secure area, diesel costs, rent since [they] can’t live in their own home, food prices are up… all while employment goes down.” (Medical Technician).

##### Increasing unaffordability of care

Further downstream, the limited resources caused by physical attacks and defunding led to increasing costs of health services, which disincentivized patients from accessing preventative care. For example, major transportation issues due to long distance and transportation costs led to inaccessibility of hospitals among civilians, exacerbating conditions that require follow-up (particularly for those living in mountainous or rural regions). Numerous private health enterprises emerged to fill this gap, but participants reported that they were not consistent, and the resulting financial pressure was then on patients themselves to pay directly for services, a grim challenge when the conflict disrupted so much employment in the area. Participants mentioned that patients frequently had to choose between two risky options: fear of being in a public hospital that is being bombed or significant financial risks from seeking care in a more expensive private facility. For example, one participant said, “Private hospitals were $50 per day which is a lot. Bombing of big or public hospitals drove people to private smaller facilities but the financial pressures drove them back to the public hospitals. …and they became afraid and began to suffer from phobia from the intense and focused bombing on the medical centers.” (Pharmacist)

#### 6) Compromised security and protection

In response to attacks on health, administrators, professionals, and patients were required to make constant calculations of security risks and plans. Many participants noted that security risks deeply impacted patients’ choice to even seek healthcare. “Even the threat of violence limits health services” said one participant. (First Responder) Several participants spoke about the deconfliction mechanism in Syria, the decision to share coordinates of hospitals with UN Member States and armed actors in a (failed) attempt to ensure respect for IHL. One of these participants said, “we did everything we had to do and we told them that those locations were medical centers, but we were regarded as enemies and then were attacked….No one… no one stopped these attacks.”(Physician). The deliberate targeting of medical facilities by the Syrian government and its allies despite consensus from health actors in northern Syria to participate in the deconfliction mechanism was particularly galling for most participants, causing deep mistrust of the UN and the international response in Syria.

##### Fortifications and contingency measures

At the facility level, insecurity resulted in adaptations and restructuring that did not always provide added protection. Fortifications of community hospitals, building hospitals outside of the community and in caves and underground, were novel adaptations to high intensity aerial attacks. While there was hope among participants that these fortifications would better protect patients and providers, this was not always the case. One participant pointed out that several fortified facilities have since been targeted and destroyed with even stronger weaponry. Beyond infrastructure modification, facilities also took steps to invest in air raid alarm programs, removing the medical emblem from their frontage, and employing armed guards to monitor the entrances to mitigate attacks.

Despite these efforts, protections were fragile and limited by the broader conflict and violence in the region. One participant explained that “We built this hospital underground, which made us feel very safe because the airstrikes that targeted the perimeter of the hospital and the hospital itself did no damage at all. However, it was not very safe because the road leading to the hospital was dangerous.” (Nurse) Transportation to or between health centers was especially fraught, “There is no public transportation. Even private transportation is dangerous. When a person moves at night, he is afraid of being sniped by a gun plane. The gun plane as soon as it sees a light at night, snipers it, regardless of whether there are children, women, or injured people.”(Administrator)

Health workers themselves also attempted to mitigate the risk to themselves and their patients. Several participants described that offices would shorten visits or send patients home quickly to mitigate risks. They also developed staggered schedules to spare staff and patients if attacks occurred.

##### Cumulative impacts on the health of the community

Collectively, strong health systems ensure that the community has a robust health care services and that population health outcomes achieve goals of “health and well-being.”[30] Participants reported that the cumulative impacts of attacks had dramatically weakened the foundation of healthcare across northern Syria, leading to significant and adverse consequences for population health outcomes including overall health behaviors, the sense well-being among the community and ultimately, mortality and morbidity (Figure 2). Participants described numerous outcomes on their patients and communities, with the most prominent themes of forced displacement, avoidant health-seeking behaviors, and a reported increase in various disease outcomes. Participants described that because of violence against healthcare and its wide health system impacts, people living in northern Syria had far fewer options to seek care, either for prevention or treatment, resulting in more illness severity, frequency and prevalence compared to before the conflict.

**Figure 2.**
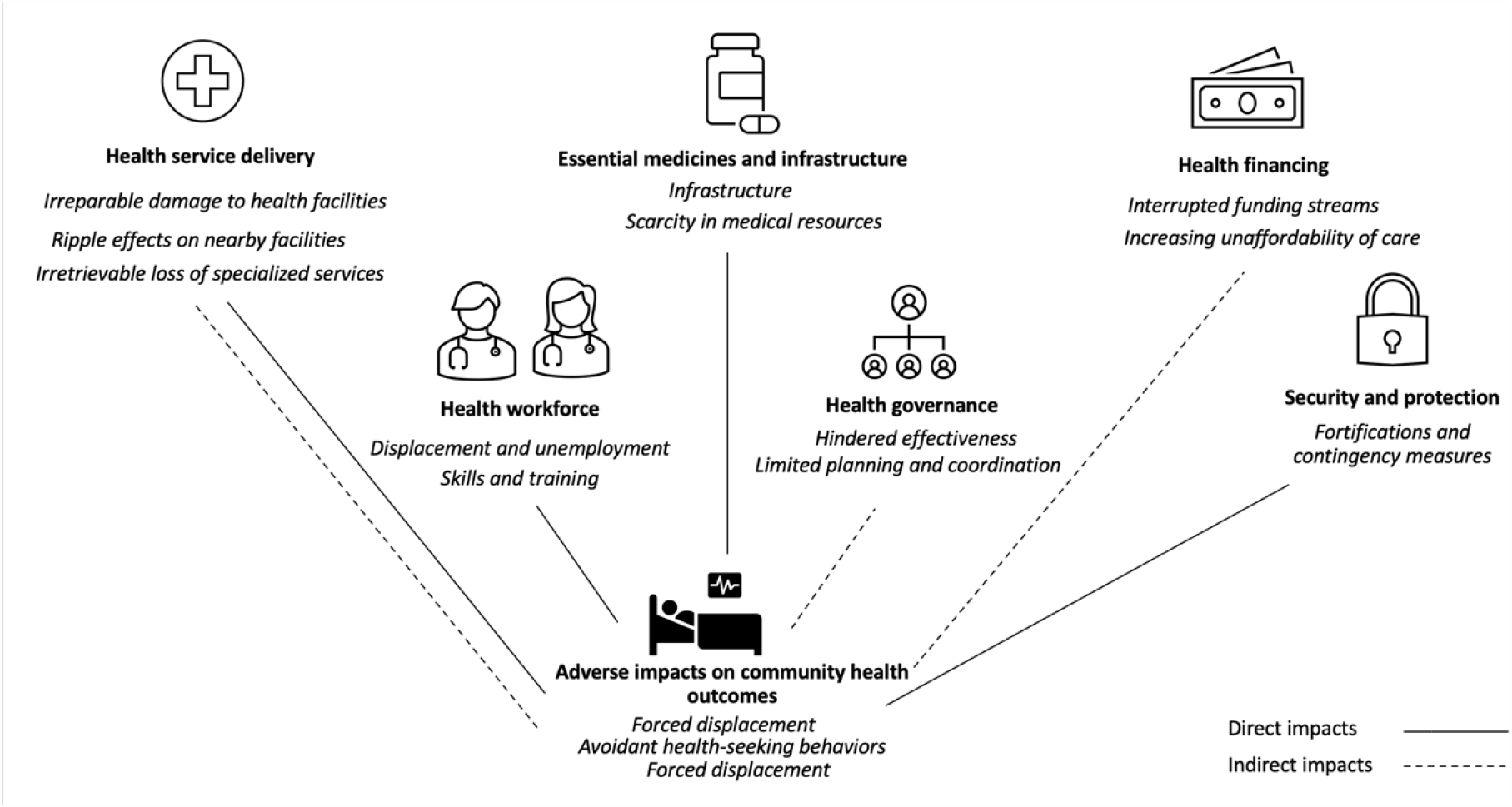
Conceptual illustration of direct and indirect health system and population impacts of attacks on health in Syria (using an adapted version of the WHO Health System Building Blocks framework)

##### Forced displacement

Protracted attacks on healthcare were reported to be a direct driver of forced population displacement. One participant explained that “After the hospital was targeted, the town became empty.” (Reception Officer) The resulting displacement also resulted in overburdening health systems not designed for mass population movements. A participant noted that “the camps put a lot of pressure on the (border) hospital which was already providing its services to people who live there.” (Technician) Those with chronic conditions also had to make difficult decisions between staying in their communities potentially risking their health or relocating nearer to health services. When a hospital closed, a person with chronic illness “had to leave the area for any medical problem, or any tiny wound would lead to death, he would not be able to get to the hospital.” (Physician)

##### Avoidant health-seeking behaviors

Civilians who stayed in northern Syria had to navigate between the practical need for health care services and the fear of getting injured or killed if they sought health services at the wrong time. One participant described his patients’ thought processes as, “it’s okay, let me feel sick a little longer [rather] than lose my life in the hospital.” (Nurse) Many participants drew a direct line from attacks on health to worsening health outcomes because of this hesitation, exacerbating their health conditions and contributing to worsening health outcomes or death.

##### Perceived increase in disease prevalence

Many participants observed increases in the prevalence of communicable disease and chronic diseases in the aftermath of attacks, possibly because diseases were left untreated in their early stages and became more severe. Chronic diseases such as heart disease and diabetes require consistent medication management, regular clinic visits, and supervision. Several participants perceived that because trauma care frequently took precedence after attacks on health, preventative and chronic care were therefore underprioritized and their severity increased. One participant noted that aid agencies and donors also influenced this change, as “they started to focus primarily on war-related injuries and casualties at the expense of the communicable and chronic diseases, which increased terrifyingly especially after 2017.” (Physician) Participants noted that attacks on health played a role in the declining workforce, displacement and lack of medications and other resources which in turn impacted communicable diseases particularly. One participant described that “several kinds of diseases have prevailed in our areas such as leishmaniasis, scabies, tuberculosis, and polio. This was all because of the lack of medical staff [to respond to these issues].”(Nurse) Participants pointed to both direct and indirect relationships between attacks on healthcare and worsening mortality and morbidity. Attacks on health were strongly associated with civilian violence in this area as well. When an attack occurred, the number of wounded and sick people would increase, just at a time when hospital services were destroyed. The incongruity between the rising need for care and the declining availability of services resulted in worse outcomes.

## IV. Discussion

This study examines the multifaceted impacts of attacks on health on the health system in northern Syria where frequent and repeated airstrikes on health facilities over the past decade have been well-documented as deliberate and strategic, a clear violation of International Humanitarian Law [2,7-16] The resultant damage to every level of the health system has cumulative effects on the population of northern Syria. This is especially pertinent in the wake of the February 2023 earthquakes, which caused further devastation to a health infrastructure already weakened by violence against healthcare.[31] As airstrikes targeting health facilities are being increasingly seen in other conflicts, including in Ukraine, Myanmar and Sudan in 2023, these impacts have unfortunate echoes elsewhere.[1-3] Insight into both the characteristics and the impacts of violence against healthcare is critical to better protecting health workers, patients and all civilians.

Analyzing our research through the lens of the WHO Health System Building Blocks provided some structure to our findings: all pillars are essential to a functioning health system and were profoundly affected by violence against healthcare.[32] Medicines and infrastructure were destroyed by the airstrikes; the lack of resources and concerns around repeat attacks made replacing them challenging. Health service delivery was disrupted both in the long and short-term, with substantial impacts on service delivery in attacked facilities as well as neighboring ones. Workforce impacts were notable not only for displacement, reductions in available workers, and task shifting, but also for paradoxical challenges with finding employment in the aftermath of an attack. With regards to governance, our findings specifically relate to how health professionals experienced governance, including the perception that health governance in northern Syria was weakened in the face of attacks, leading to a fragmentation of service provision.[15, 34-36] Other research has considered the impacts of attacks on healthcare on governance in greater detail [38, 42-44]. Financing disruptions have also resulted in some misalignment between local needs and international donors with unstable funding streams.[37]

Several cross-cutting findings also emerged. First, violence against one component of the health system had impacts on others. For example, governance challenges made the rebuilding of the workforce more difficult. [38,39] Medicines were destroyed by the airstrikes and the lack of financing made replacing them challenging. Health infrastructure was disrupted both in the long and short-term, with substantial impacts on service delivery in attacked facilities as well as neighboring ones. Also, impacts were compounded over time, as we hypothesized in conceptual work [33] For instance, health workers noted that the repeated nature of attacks on health in Syria over prolonged periods resulted in impacts far greater than each individual attack. Third, the study exposed several complex causal pathways between the attack and the outcome, suggesting that there are multiple opportunities to intervene and ameliorate population health impacts. For example, the growing burden of chronic illness resulted from several layered processes including donors sometimes prioritizing funding for trauma care (resulting in fewer chronic care services) and patients who were afraid of violence against healthcare waiting longer to seek care (an adaptive response which has been identified in previous studies) and task shifting of health workers towards emergency care.[7-10] Interventions at any of these steps in the cascade could mitigate the burden of disease. This study also found that knock-on effects reached far beyond the health outcomes, with impacts on population displacement as well as social cohesion. Finally, the findings touch on the concern that while impacts were severe across the conflict, they were not homogenous. Often, the most vulnerable communities (children, chronically or severely ill, rural communities) suffered disproportionately, particularly those that had experienced several displacements or continued to be internally displaced. [[1-3,7-10,33]

Our research adds to growing literature on the impacts of conflict and targeted attacks on health. For example, our findings on increasingly siloed health programs and fragmented leadership are also explored by Alkhalil et. al., who note that the multiple Health Directorates were established soon after the withdrawal of the Ministry of Health.[37] Our study underscores the Douedari et. al. finding that a key constraint of health system reconstruction is the ongoing targeting of health facilities and health workers.[40] This has been compounded by the exodus of health workers with many of those leaving among the more senior, experienced and more able to leave due to having recognized degrees and sought-after experience.[41] While the interviews only gave a glimpse of health worker perspectives on governance issues, they support extensive literature on the complexities of northern Syria’s governance challenges including the evolution and performance of the local health directorates, the competition between humanitarian structures and the directorates and the refusal of Syria’s health ministry to engage.[42-44]

Although deepening our understanding of the impacts of violence against healthcare on health systems from the perspectives of affected health workers is critical, it is essential to operationalize this knowledge with more targeted and concrete strategies to engage local health workers, support them, seek accountability, and protect them from attacks. The health workers we interviewed were able to share their perspectives not only as direct targets but also as wider members of the community, underscoring a need for better protections and support. Of importance is that non-physicians and women health workers must be equally centered in these conversations. The health workforce remains the foundation of the health system; polices and frameworks which protect and safeguard their physical, psychological, and financial well-being are essential. Steps to concretely achieve this goal may include encouraging existing, local, quasi-governmental bodies and funders to support formal arrangements for salary protection in the wake of attacks or insurance should they be injured or killed, supporting training initiatives such as the Syrian Board of Medical Specialties, and continuing to have health workers engaged in broader decision making regarding the health system.[19, 37,45,46] For the international community, supporting health systems in conflict-affected Syria should involve stabilizing funding streams for equipment, medicines and infrastructure that prioritize local needs, supporting local governance, ensuring long-term and sustainable care delivery systems and developing sustainable pipelines for training new professionals across the various professions. [19]

The health system impacts resulting from attacks on healthcare that have been explored in this study are complex and to an extent entrenched; they will last decades, if not generations. Knowing this, the prevention of attacks must be strengthened. Such attacks are violations of International Humanitarian Law. Given current paralysis in accountability mechanisms, states’ abdication of responsibility for implementation of UN Security Council Resolution 2286, and the scale of such attacks, insights from this and similar work are essential to stimulate the political will to prevent violence against health, hold perpetrators to account and to mitigate their impacts.[47]. Our work emphasizes the need for research beyond counting the number of attacks and the quantitative extent of damage resulting from violence against healthcare. Taking a health systems approach and exploring the structural impacts through the voices of health workers directly affected (and, where possible civilians and patients) is essential to build our understanding of the multiple, compounding, and intersecting aspects of this topic. This is increasingly relevant where current, active conflicts in Myanmar, Sudan, and Ukraine, among others, necessitate this approach to support a nuanced and contextualized understanding of the impacts [25].

Further research could include more diverse voices, including community members and health workers both from Syria as well as the many other contexts where healthcare is targeted.[31,34,36] Next steps may also consider mixed-method or quantitative analysis of the health system impacts identified in this study; a more granular analysis could identify differential impacts based on the type and severity of attack on specific communities and their respective health systems.

### Limitations

As in all studies involving interviewers and interviewees, there is a risk of recall and reporting bias, or bias introduced by the researchers. The sampling approach and including only health workers and those living in primarily northern Syria, limited our understanding of the perspectives of the local community, other aid workers, leadership and those not directly experiencing violence. This may particularly limit analyses of some health system components such as governance and financing where health workers may not play a direct role and administrators and other stakeholders may be able to give a fuller picture. We interviewed health workers who remained and continued working in Syria and did not include the perspectives and experiences of those who left or stopped working in healthcare. Though we had a diverse set of participants, including students, technicians, and mid-level providers, we sampled fewer female health workers than we hoped, despite active efforts to recruit female participants and ensure a female interviewer was available. This was largely due to hesitations expressed by female health workers to participate in a research study citing safety and confidentiality. We hope future studies expand the sampling frame, include more women, and expand on the understanding of the many impacts of conflict and targeted violence.

### Conclusion

This study provides compelling evidence of the profound and far-reaching consequences of violence against healthcare in Syria and its multilateral impacts on the health system. Our findings highlight the compounding, cumulative nature of these impacts, which have reverberating effects on the health of the communities served by directly and indirectly affected health facilities. It is crucial for efforts to protect health in conflict settings to address not only the attacks themselves but also the devastating repercussions they have on vulnerable individuals and systems, emphasizing the need for accountability, resource prioritization, and prevention of attacks.

## Data Availability

The qualitative datasets presented in this manuscript are not readily available because of the sensitive nature of the data. There are ethical requirements that stipulate that qualitative data cannot be shared.

## V. Declarations

### Ethics Statement

Ethical approval for this study was granted by the Human Research Protection Program at the University of California, Berkeley (Protocol ID# 2020-03-13069) and reviewed by the Idlib Health Directorate. Formal consent was obtained from all participants in this study.

### Consent for publication

Not applicable; all data are de-identified. In order to ensure confidentiality, the interview was completely anonymous, and the transcribed interviews were de-identified, such that no identifying information could be detected when using verbatim quotes.

### Competing interests

The authors have no competing issues to declare. We confirm that all authors have approved the manuscript for submission.

### Funding

Funding for this project and all authors was provided by the Research for Health in Humanitarian Crises (R2HC, https://www.elrha.org/programme/research-for-health-in-humanitarian-crises/) program at ELRHA (Grant #35189, PI RH). The funders had no role in study design, data collection and analysis, decision to publish, or preparation of the manuscript.

### Authors’ contributions

RH, AA, LR and HT designed the study and conceptualized the methodology. DR, HT and RH developed the methodology. DR and AR conducted the interviews. RW and HA translated and transcribed all the interview manuscripts. DR, AR, HT, AA and RH coded and analyzed the data. RH and DR wrote the initial manuscript draft. All authors read, revised and approved the final manuscript.

## Acknowledgements

We thank Syrian American Medical Society, Assistance Coordination Unit, the Researching Impacts of Attacks on Health Consortium and Physicians for Human Rights their invaluable contributions to this research, including Houssam Alnahhas and Christian DeVos at PHR, Larissa Fast and Christina Wille with RIAH, Qasim Yagizi, Mohamad Rami Kawas and Aya Aksh at ACU, Ahmad Tarakji and Randa Loutfi at SAMS. We thank the Idlib Health Directorate for supporting this study. We would also like to thank the dedicated health workers across northern Syria who generously shared their experiences and insights with us.

## Notes

### Competing Interest Statement

The authors have declared no competing interest.

### Author Declarations

The University of California, Berkeley Human Subjects Protection Program reviewed and approved this study (Protocol 2020-03-13069). The researchers also consulted with and received approval from the Idlib Health Directorate and other local leadership in lieu of a formal ethical review board in Syria. Comprehensive informed consent procedures took place at the start of each interview. Participants were able to terminate the interview at any time and to withdraw their participation until the time at which analysis was started. Given that some topics covered may be sensitive or cause distress, a reference to a mental health professional was available should this be needed by the participants.

